# Effects of Aging, Hearing Loss, and Co-Activation on the Middle Ear Muscle Reflex and Medial Olivocochlear Reflex

**DOI:** 10.64898/2026.04.27.26351829

**Authors:** Pauline Devolder, François Deloche, Morgan Thienpont, Hannah Keppler, Sarah Verhulst

**Affiliations:** Hearing Technology @ iBME, Department of Information Technology, Ghent University, Zwijnaarde, Belgium; Department of Rehabilitation Sciences, Ghent University, Ghent, Belgium; Department of Otorhinolaryngology, Head and Neck Surgery, Ghent University Hospital, Ghent, Belgium

**Keywords:** middle ear muscle reflex, medial olivocochlear reflex, efferent pathways, aging, sensorineural hearing loss

## Abstract

The middle ear muscle reflex (MEMR) and medial olivocochlear reflex (MOCR) are increasingly studied for their role in suprathreshold auditory processing. However, recording these reflexes in humans is potentially complicated by age-related (sub)clinical hearing loss and co-activation. This study investigates the influence of age-related sensorineural and audiometric hearing loss, and how MEMR activation contaminates MOCR recordings.

Three test groups were included: young normal-hearing adults, middle-aged normal-hearing adults, and middle-aged adults with hearing loss. The sensorineural auditory status was assessed using distortion-product otoacoustic emissions (DPOAEs) and envelope following responses. MEMR recordings were evaluated using wideband stimuli and MOCR was recorded at elicitor levels of 60 and 75 dB to evaluate MEMR co-activation.

MEMR amplitude, but not threshold, was related to age; however, this effect was significantly reduced after correcting for DPOAE and extended high-frequency thresholds. Associations between MOCR strength and age or hearing-related measures were mixed and reliable recordings were challenging in participants with hearing loss due to poor OAE baselines. MEMR co-activation was detectable in the click response and could alter estimated MOCR-induced suppression.

These findings suggest that MEMR amplitude may be a marker of subclinical cochlear damage, while MOCR sensitivity to hearing status appears more closely tied to audiometric than subclinical sensorineural status. Clinical measurements can be improved using broadband stimuli, accounting for cochlear damage, and defining criteria for reflex co-activation.

## Introduction

Auditory reflexes have gained increasing attention in hearing research due to their possible relation to suprathreshold sound encoding. The middle ear muscle reflex (MEMR) has been suggested as potential indicator of cochlear synaptopathy (Bharadwaj et al., 2019; Mepani et al., 2020; Valero et al., 2018; Wojtczak, Beim, & Oxenham, 2017), while the medial olivocochlear reflex (MOCR) is thought to play a role in facilitating speech perception in noisy environments (Abdala et al., 2014; Guinan Jr et al., 2003; Lopez-Poveda, 2018). However, measuring and isolating these reflex pathways in humans remains challenging due to (1) confounding factors such as audiometric thresholds and “hidden” sensorineural hearing deficits that are not observed via tonal audiometry, and (2) co-activation between the two reflexes.

Studies have shown that both MEMR and MOCR activity declines with age (Abdala et al., 2014; Feeney, Keefe, & Marryott, 2003; Keppler et al., 2010; Parthasarathy, 2001). However, audiometric and sensorineural hearing loss may influence these measurements and could interact with age-related findings. These auditory reflexes rely on afferent (ascending) and efferent (descending) neural pathways: afferent inputs carry signals from the cochlea to the brainstem or higher, and efferent projections return from the brainstem to the auditory periphery. Consequently, depending on the specific pathways involved, peripheral sensorineural damage could influence reflex measurements. In cases where this is desired, for instance when using MEMR as a marker of cochlear synaptopathy, it is important to ensure that we can isolate this factor without the influence of other (sub)clinical hearing damage, such as outer-hair-cell loss. Alternatively, if we want to use a reflex measurement to isolate the efferent pathways, the influence of subclinical hearing loss is undesirable.

The MEMR pathway originates at the inner hair cells, travels through the cochlear nucleus and superior olivary complex, and reaches the facial motor neurons. These neurons send efferent fibers via the facial nerve to the stapes muscle that stiffens the ossicular chain. The exact neural circuit is complex and not yet fully described, see Mukerji, Windsor and Lee (2010) for more details. Because this pathway includes the inner hair cells, auditory nerve, facial nerve, and middle ear, MEMR measurements could theoretically be affected by middle-ear abnormalities, facial nerve pathology, auditory neuropathy, cochlear synaptopathy, or inner hair cell damage. Functionally, it has been suggested that the MEMR may protect the ear from acoustic injury and could optimize auditory perception during loud sounds or vocalization (Borg, 1984; Borg, Nilsson, & Engström, 1983; Liberman & Guinan Jr, 1998). The reflex is typically elicited at suprathreshold levels, causing stapedial muscle contraction that stiffen the ossicular chain, alter middle-ear impedance, and bilaterally attenuate low-frequency cochlear input (Kobler et al., 1992; Møller, 1974). MEMR measurements are generally performed with high-intensity probe levels (90 dB peSPL) to asses changes in ear-canal reflections after eliciting the reflex. The influence of cochlear gain, and thus outer-hair-cell damage, should be negligible at these high stimulation levels.

The MOCR involves afferent projections from inner hair cells to the cochlear nuclei and superior olivary complex, where they synapse onto efferent fibers that descend via the auditory nerve to the outer hair cells, reducing their electromotility (Brown et al., 2013). Similar to the MEMR, the MOCR’s exact pathway is not fully understood yet, as well as potential higher pathway contributions, e.g. at the level of the inferior colliculus (Mulders & Robertson, 2000). MOCR measurements could be sensitive to damage affecting outer hair cells, efferent pathways, auditory neuropathy, cochlear synapses, or inner hair cells. The MOCR is typically activated at lower elicitor stimulation levels than the MEMR (Guinan Jr et al., 2003; Marks & Siegel, 2017) and reduces outer-hair-cell motility, thereby decreasing cochlear amplification (Guinan Jr et al., 2003). While its exact function is still uncertain, the MOCR is believed to enhance the detection of sound in noise (Abdala et al., 2014; Guinan Jr et al., 2003; Liberman & Guinan Jr, 1998; Lopez-Poveda, 2018; Maison & Liberman, 2000). There is increasing interest regarding its role in auditory processing (Boothalingam et al., 2019; Boothalingam et al., 2016; Guinan Jr, 2018; Mertes, 2024). MOCR measurements are generally performed with low to moderate probe levels (55 dB peSPL when click-evoked) to asses changes in otoacoustic emissions after eliciting the reflex.

Over the past decades, considerable research has focused on optimizing protocols to evaluate these reflexes. First, a consistent finding for MEMR is that wideband measurements provide more reliable and sensitive measurements than the traditional 226 Hz clinical stapes reflex testing (Bramhall et al., 2025; Feeney & Keefe, 1999; Feeney, Schairer, & Putterman, 2023; Schairer et al., 2007). Wideband approaches use a click or chirp as probe stimulus instead of a low-frequency tone, thereby capturing changes in ear-canal reflectance across multiple frequencies. This wide frequency information is advantageous because individual differences in ear-canal acoustics and impedance can influence compliance measures (Feeney & Keefe, 1999). Compared with conventional 226-Hz probe tones, wideband probe stimuli evoke reflexes at lower intensities, providing more sensitive measurements, richer information on reflex growth, and less susceptibility to ceiling effects (Feeney & Keefe, 2001; Mepani et al., 2020).

Secondly, it is known that MEMR activation can contaminate MOCR measurements, particularly in OAE-based paradigms (Guinan Jr et al., 2003). Since MEMR contraction alters middle-ear impedance in a frequency-dependent manner, it can either reduce or enhance the stimulus level reaching the cochlea, depending on the spectral content of the stimulus, and thereby alter MOCR responses. To minimise this effect, MOCR measurements are typically performed with low-level elicitor noise (often around 60 dB SPL), which is assumed to fall below the MEMR threshold while still evoking measurable OAE suppression. It has been suggested that changes of the stimulus waveform recorded in the ear canal could be an indicator of this MEMR involvement (e.g., in Boothalingam et al. (2019)). Despite these precautions, it remains unclear to what extent MEMR activation occurs during MOCR testing at a 60-dB elicitor level, how this contamination can be estimated and whether the effects are most prominent at low frequencies, where MEMR-induced impedance changes typically occur.

This study aims to improve interpretation of MEMR and MOCR measurements in clinical and scientific settings to further study the diagnostic value of these reflex measurements. We investigated the following three research questions:

First, how are MEMR and MOCR measurements affected by age and audiometric hearing loss? MEMR and MOCR were measured in three participant groups: young adults with normal hearing (yNH), older middle-aged adults with normal hearing (oNH), and older middle-aged adults with audiometric hearing loss (oHI). Comparisons between yNH and oNH assessed age-related effects independent of audiometric hearing loss, while comparisons between oNH and oHI examined the impact of audiometric hearing loss.

Second, to what extent are age-and audiometry-related effects on MEMR and MOCR explained by afferent sensorineural hearing deficits and audiometric thresholds? Sensorineural damage was evaluated using distortion product otoacoustic emissions (DPOAE; outer-hair-cell function) and the envelope following response (EFR; suggested marker of cochlear synaptopathy). In particular, a better understanding of the relationship between EFR and MEMR could clarify their potential role as markers of cochlear synaptopathy.

Third, to what extent does MEMR activation influence MOCR measurements, and can co-activation be observed during MOCR testing? Both MEMR and MOCR were assessed using similar click and noise stimuli. MOCR measurements were conducted at a commonly used elicitor level (60 dB SPL) below the typical MEMR threshold and at a higher level (75 dB SPL) just above most participants’ individual MEMR threshold. MEMR thresholds were measured using the standard approach with an intensity-growth paradigm from 40 to 90 dB SPL, in order to define the threshold. This approach allowed us to link MEMR activation to MOCR responses and quantify potential co-activation effects.

## Methods

### Participants

To investigate the effects of age and audiometric hearing loss on MEMR and MOCR measurements, we considered three test groups:

1. 20 young adults with clinically normal pure-tone audiograms (yNH), age 21 to 35 years (M = 27.2; SD = 3.62), 10 male and 10 female subjects
2. 14 middle-aged adults with clinically normal pure-tone audiograms (oNH), age 46 to 62 years (M = 53.4; SD = 5.20), 6 male and 8 female subjects
3. 17 middle-aged adults with clinical hearing loss (oHI), age 47 to 65 years (M = 55.1; SD = 6.13), 9 male and 8 female subjects

Normal hearing was defined by hearing thresholds ≤ 20 dB HL at all frequencies on conventional audiometry: 125, 250, 500, 1000, 2000, 3000, 4000, 6000 and 8000 Hz. Only ears with type A (n = 46) or As (n = 5) tympanograms were included to reduce the likelihood of conductive hearing loss. Only one ear, with the best audiometric thresholds (lowest maximum threshold or mean threshold at 0.5, 1 and 2 kHz) was tested in the protocol.

This research was approved by the ethical committee at Ghent University (ONZ-2022-0352 E01). Participants signed an informed consent before the experiment.

### Audiometric and sensorineural hearing status

Tympanometry was performed using the Sentiero Tymp Diagnostic (Path Medical). A probe containing a speaker and microphone was placed in the participant’s ear canal with an air-sealing probe tip. A probe tone of 85 dB SPL at 226 Hz was used to measure impedance during the air pressure shift. Audiometric thresholds were measured using the modified Hughson-Westlake method. Audiometry was carried out in a double-walled booth using the Equinox Interacoustics audiometer and RadioEar DD450 headphones. Stimuli were presented on the (half)-octave frequencies (0.125, 0.250, 0.5, 1, 2, 3, 4, 6, and 8 kHz) and the extended high frequencies (EHF;10, 12.5, 14 and 16 kHz).

To measure outer-hair-cell integrity, DPOAE testing was conducted using the Universal Smart Box Intelligent Hearing Systems and SmartDPOAE software. DPOAE stimuli were presented through an Etymotic Research 10D probe. Two tones with a different frequency were presented at a fixed primary frequency ratio f2/f1 = 1.22. Responses were recorded using a primary frequency sweep (DP-gram) from 0.5 to 8 kHz at stimulus levels of L1 =65 dB SPL and L2 =55 dB SPL during 32 sweeps. When the measured amplitude at a specific frequency was smaller than the noise floor, its value was changed to that of the noise floor to remove extreme, unreliable negative values.

To establish a possible link with cochlear synapse integrity, EFRs were measured using auditory evoked potential recordings as described in Garrett et al. (2025b). EFR’s are suggested to be a potential measure of cochlear synapse damage based on animal studies (Garrett et al., 2025a; Kujawa & Liberman, 2015; Liberman & Kujawa, 2017; Parthasarathy & Kujawa, 2018), human computational modelling (Vasilkov & Verhulst, 2019) and age-related magnitude reductions in humans with normal audiograms (Bramhall et al., 2019; Garrett et al., 2025b). The EFR was elicited using a rectangular amplitude-modulated (RAM) tone with a carrier frequency of 4000 Hz and a modulation frequency of 110 Hz. Stimuli were presented in alternating polarity at 70 dB SPL, with a duration of 490 ms and a presentation rate of 2 Hz. Each recording consisted of 800 sweeps. Signals were band-pass filtered (30–1500 Hz), epoched, baseline corrected, and analysed using a bootstrapping approach to estimate the noise floor (Van Der Biest et al., 2023; Vasilkov et al., 2021). The skin was prepared with NuPrep gel to minimize electrode–skin impedance and disposable electrodes (Ambu Neuroline) were positioned: the noninverting electrode (+) at Fz, as close as possible to Cz; the inverting electrodes (–) on the ipsilateral and contralateral earlobes (A1, A2); and the ground electrode on the nose. Signals were recorded using the Universal Smart Box (Intelligent Hearing Systems) and the SmartEP Continuous Acquisition Module (SEPCAM) software. Participants were seated comfortably in a reclined chair and instructed to remain still and relaxed. All lights and electrical devices (e.g., phones, computers) were turned off during testing to minimize electrical interference.

### Middle ear muscle reflex

#### Conventional 226 Hz acoustic reflex measurement

The Sentiero Tymp diagnostic (Path Medical) was used to measure clinical acoustic reflexes, performed immediately after tympanometry. A 226-Hz probe tone at 85 dB SPL was used in conjunction with a reflex-activating stimulus presented to the ipsilateral ear at tympanometric peak pressure (see Figure 3a). These stimuli were 500 Hz,1000 Hz, 2000 Hz, 3000 Hz and 4000 Hz tones, and broadband (BBN), low pass filtered (LPN; 891-1120 Hz) and high pass filtered noise (HPN; 3560-4490 Hz), see Figure 1. The intensity changed in increments of 5dB HL starting at 60dB HL to a maximum 105dB HL for tones and 95 dB HL for noise.

**Figure 1:**
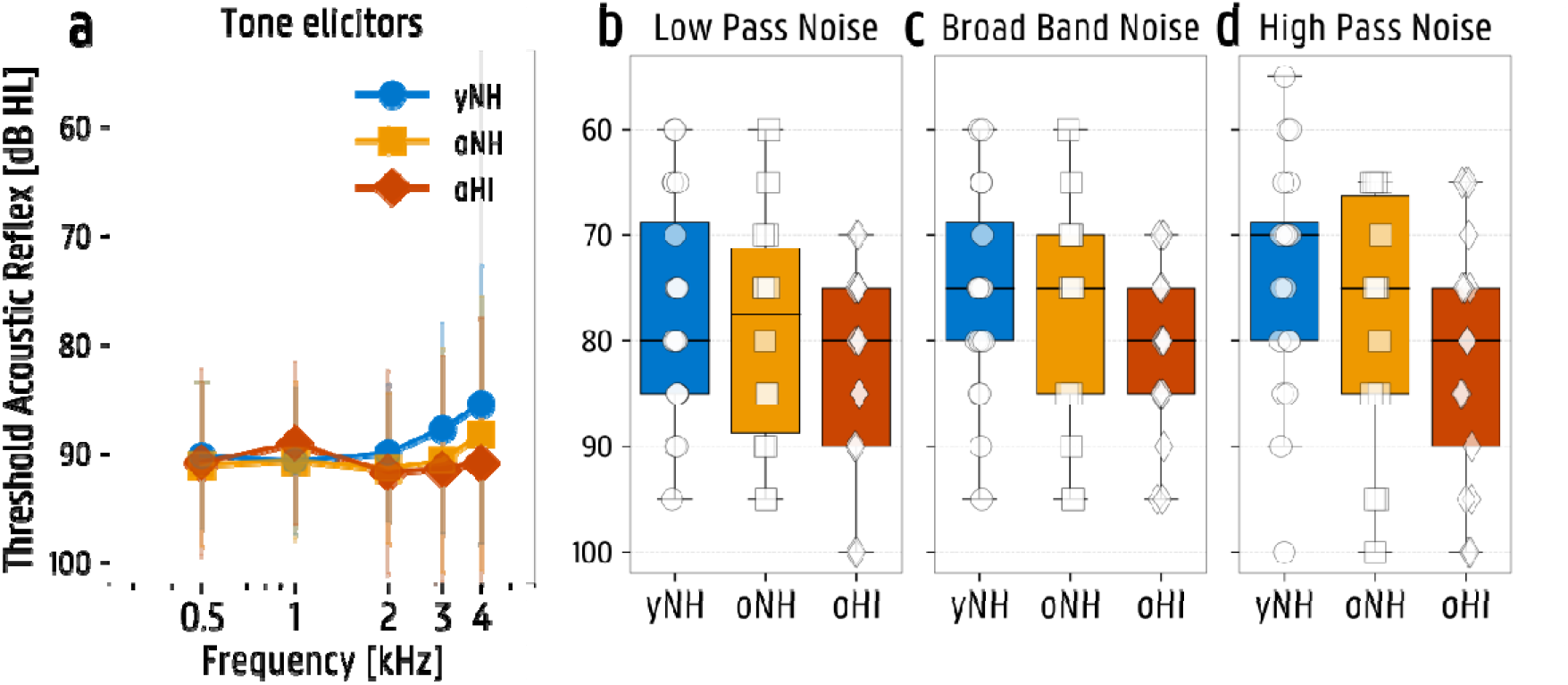
Acoustic reflex thresholds for the three test groups; yNH = young normal hearing (N=19), oNH = older normal hearing (N=11), oHI = older hearing impaired (N=13). (a) Average reflex thresholds for tone elicitors at different frequencies. (b, c, d) Boxplots of the reflex thresholds using a low pass noise elicitor broad band noise elicitor and high pass noise elicitor. After Bonferroni correction: no significant differences.

#### Wideband click-evoked MEMR

Participants were seated in a sound-attenuating booth and were instructed to remain awake and sit still during testing. Measurements were conducted using an Etymotic Research ER-10X Extended Bandwidth Research Probe System connected to an RME Fireface UCX sound card for stimulus delivery and acquisition. Probes were mounted on a microphone stand to prevent them from falling out of the ear. Stimuli were generated in MATLAB and calibration was performed using an ear simulator (Brüel & Kjaer Type 4157) and sound level meter (Brüel & Kjaer Type 2606). Clicks were peSPL calibrated to a 1kHz-pure tone using an oscilloscope (Keysight InfiniVision DSOX2022A) connected to a preamplifier (Brüel & Kjaer Type 5935).

For measuring MEMRs, we adopted the protocol of Bharadwaj et al. (2022). Wideband MEMRs were measured ipsilaterally, contralaterally, and bilaterally using 94 µs clicks at 90 dB peSPL as probe stimuli. Broadband white noise elicitors (0.5-8.5 kHz, flat excitation pattern) were presented at levels from 40 to 90 dB SPL in 5 dB steps. Each MEMR trial (see Figure 3b) consisted of a train of seven probe clicks alternating with 120 ms ipsilateral noise bursts. The interval between the click and elicitor onset was 28 ms, and the gap between elicitor offset and the subsequent click was 14 ms. The level series was repeated 16 times per ear with a 1.5 s intertrial interval to allow immittance recovery. Because participant movement occasionally affected probe placement, we removed unreliable trials before curve fitting. Outlier trials were excluded when exceeding two standard deviations from the mean of the 16 trials.

Ear canal sound level was measured within the first 20 ms of each click response (indicated in Figure 3c) and converted to changes in absorbed power. The sound level spectrum was estimated using a multitaper method. As in Bharadwaj et al. (2022), growth functions were baseline-corrected by subtracting the mean of the responses at 40, 45 and 50 dB SPL and fitted with a three-parameter sigmoid function. The MEMR threshold was defined as the elicitor level at which the fitted growth function reached 0.02 dB above the 40-50 dB SPL mean baseline. Measurements were accepted only when the goodness of fit of the sigmoid function and response characteristics met predefined criteria. In case of a bad sigmoid fit (R² < 0.6) the measurement was excluded for that subject, except when this bad fit was due to extremely shallow growth (maximal slope < 0.001), which were considered absent responses. Computed thresholds above the maximum elicitor level (90 dB SPL) were reassigned to 95 dB SPL, as were curves with extremely shallow growth (maximal slope < 0.001), which were considered absent responses. The number of excluded measurements is summarized in Table 1.

**Table 1:** Percentage (n/N) of subjects excluded from ipsilateral and contralateral middle ear muscle reflex (MEMR) analyses and of subjects with absent MEMR responses at 90 dB SPL, for whom the threshold was set to 95 dB SPL. yNH = young normal hearing; oNH = older normal hearing; oHI = older hearing impaired.

|  | Excluded subjects |  | No response found at 90 dB SPL<br>(shallow growth) |  |
| --- | --- | --- | --- | --- |
|  | Ipsilateral MEMR | Contralateral MEMR | Ipsilateral MEMR | Contralateral MEMR |
| yNH | 10% (2/20) | 0% (0/20) | 5.5% (1/18) | 10% (2/20) |
| oNH | 7.1% (1/14) | 14.3% (2/14) | 7.7% (1/13) | 8.3% (1/12) |
| oHI | 11.8 (2/17) | 35.3% (6/17) | 20% (3/15) | 45.5% (5/11) |

To quantify the advantages of the click-evoked wideband MEMR technique over the conventional 226-Hz evoked MEMR technique, we compared MEMR threshold sensitivity. Figure 2a shows the difference between the 226-Hz MEMR with a 1000 Hz elicitor, commonly used in clinical practice, and the click MEMR with a broadband noise elicitor. Within the yNH group, 18 subjects (90%) had lower (better) thresholds with the click method (positive difference). Within the oNH and oHI group this corresponded to 12 (85.7%) and 15 (88.2%) subjects respectively. Figure 2b shows the difference between the 226-Hz MEMR with a 1000 Hz elicitor and the 226-Hz MEMR with a broadband noise elicitor. All subjects (100%) had lower thresholds with the broadband noise condition. Figure 2c shows the difference between the 226-Hz MEMR with a broadband noise elicitor and the click MEMR with a broadband noise elicitor. In the yNH group, 13 subjects (65%) had better thresholds with the click MEMR. Within the oNH and oHI group this corresponded to 6 (42.9%) and 10 (58.8%) subjects respectively. These results suggest that the elicitor-based stimulus adaptations were consistently more advantageous than probe-based stimulus adaptations. For the next analyses, wideband MEMR data is used, involving clicks as probe-stimuli and broadband noise as elicitor.

**Figure 2:**
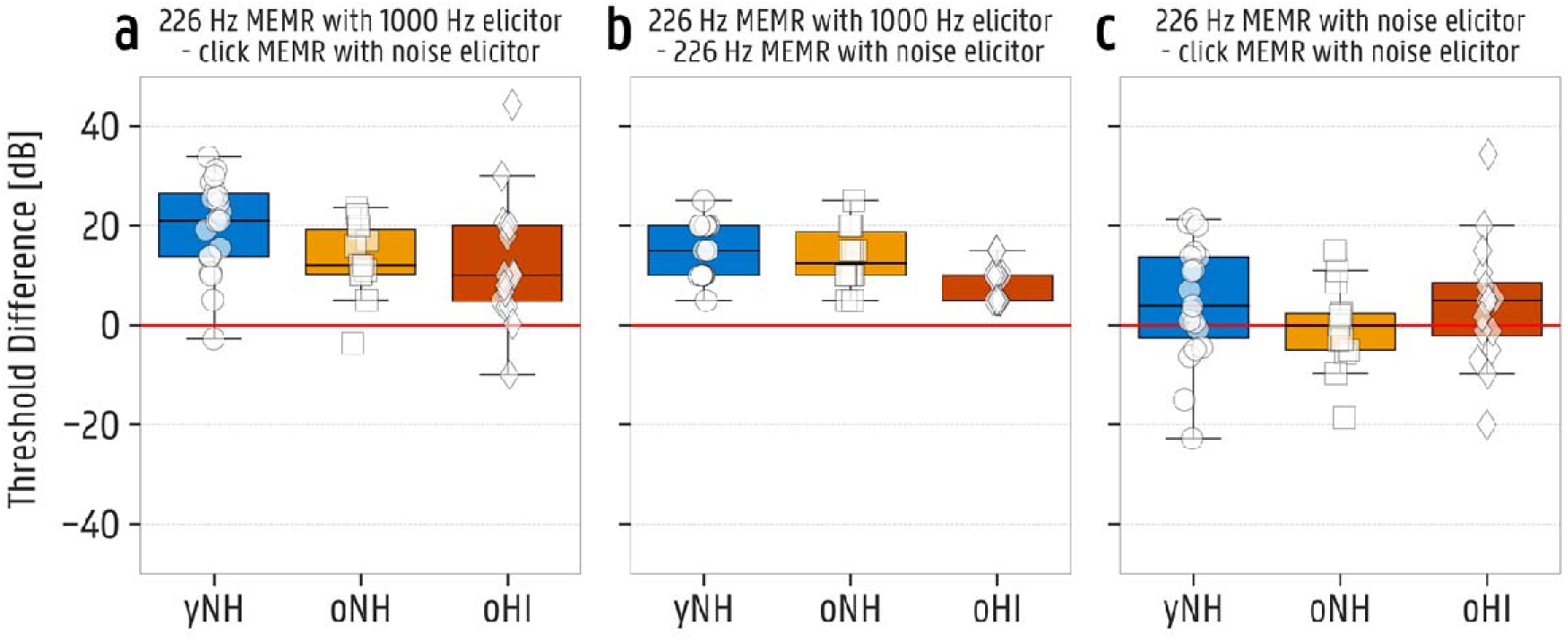
Difference between thresholds of different MEMR techniques for the three test groups: yNH = young normal hearing (N=19), oNH = older normal hearing (N=11), oHI = older hearing impaired (N=13). (a) Boxplots of the threshold difference calculated as the 226 Hz middle ear muscle reflex (MEMR) with a 1000 Hz elicitor minus the click MEMR with a broadband noise elicitor. (b) Boxplots of the threshold difference calculated as the 226 Hz MEMR with a 1000 Hz elicitor minus the 226 Hz MEMR with a broadband noise elicitor. (c) Boxplots of the threshold difference calculated as the 226 Hz MEMR with a broadband noise elicitor minus the click MEMR with a broadband noise elicitor.

### Medial olivocochlear reflex

The same setup as for the wideband MEMR measurement was used, but with modifications in the stimulus paradigm and intensities. Instead of assessing changes in ear-canal reflections, MOCR recordings measure changes in cochlear amplification via otoacoustic emissions (OAEs) (Guinan Jr et al., 2003). Various OAE types, including transient-evoked (TEOAEs), distortion-product (DPOAEs) and stimulus-frequency (SFOAEs), can serve as probe stimuli of the MOCR (Goodman, Boothalingam, & Lichtenhan, 2021; Guinan Jr et al., 2003; Jedrzejczak et al., 2024; Mertes & Goodman, 2016). Click-evoked otoacoustic emissions (CEOAEs) were measured ipsilaterally, contralaterally, and bilaterally using a forward-masking paradigm adapted from Boothalingam et al. (2019) (see Figure 3d). Probe clicks (93.75 µs, 55 dB peSPL) were presented in 96-ms blocks at a rate of 41.67 Hz. As for the wideband MEMR measurement, noise served as elicitor and was presented at a fixed level. We adopted the same broadband noise elicitor as for the MEMR, but at a level of 60 dB SPL and with a duration of 478 ms. Each trial consisted of five sequential blocks: baseline, ipsilateral noise, contralateral noise, bilateral noise, and a second baseline, each time followed by a click train. A shortened visualisation of the ipsilateral measurement can be seen in Figure 3d, the complete stimulus protocol is described in Boothalingam et al. (2019). A total of 400 trials were collected, resulting in 1600 click epochs per condition (3200 for baseline).

**Figure 3:**
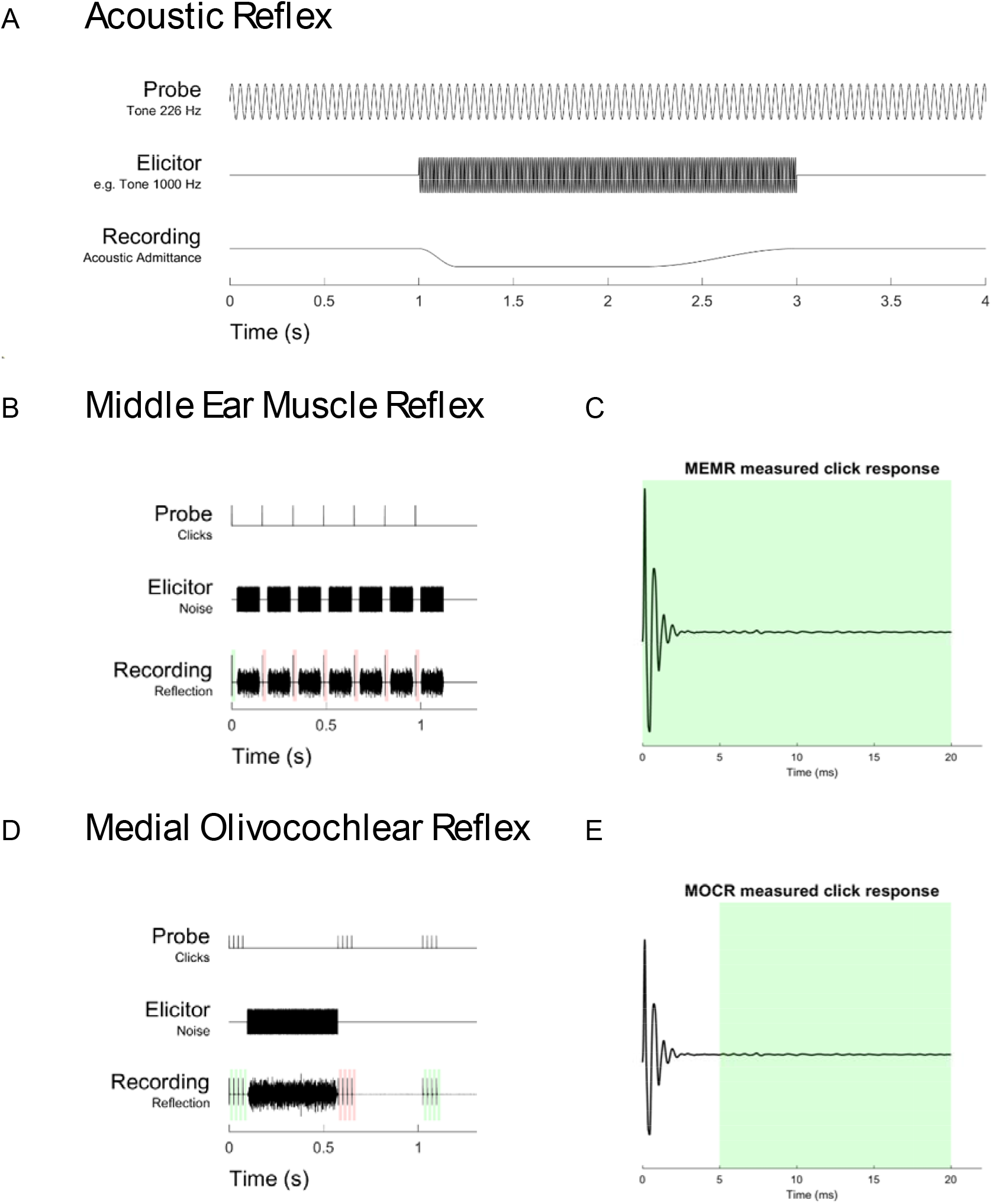
Visualisation of the probe stimulus, elicitor stimulus and recording of the reflex measurements. (A) Stimuli for the conventional 226 Hz tone-based acoustic reflex measurement. (B) Stimuli for the wideband middle ear muscle (MEMR) reflex measurement. (C) Visualisation of a measured click response with the MEMR analysis window indicated in green. (D) Stimuli for the wideband medial olivocochlear reflex (MOCR) measurement. (E) Visualisation of a measured click response with the MOCR analysis window indicated in green.

CEOAE responses (5-20 ms post-click, indicated in Figure 3e) were bandpass filtered (0.6-6 kHz, zero-phase fourth-order Butterworth). Epochs exceeding 2 SDs above mean RMS amplitude were rejected. Responses were separated into odd-and even-numbered buffers. After averaging, the CEOAE magnitude was estimated from the mean of the two buffers, and the noise from their difference divided by sqrt 2.. If the reproducibility of the baseline CEOAE (odd-even correlation of both baseline buffers) was less than 80%, the measurement for that subject was excluded. Figure 4a indicates that this was especially the case in the oHI group. Eight measurements (47.1%) were excluded within this group, while only 4 (28.6%) and 3 (15%) subjects were excluded in the yNH and oNH groups respectively. The wideband baseline SNRs are shown in Figure 4b, indicating that the noisiest measurements had already been excluded by the reproducibility criterion. For spectral analysis, frequency bins were retained only if their baseline CEOAE magnitude exceeded the noise floor by ≥12 dB. Unlike the procedure described by Boothalingam et al. (2019), frequency bins showing enhancement (i.e., higher CEOAE magnitude in the elicited than baseline condition) were not discarded. Instead, all retained bins contributed to the final ΔCEOAE metric through a weighting scheme. Specifically, let P_1_(f) and P_2_(f) be the power spectrum of the baseline and elicited responses. We then define the weights as *w*(*f*) = P_1_(*f*)/Σ_i_P_1_(*f_i_*), the frequency-specific Delta as 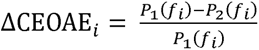 and ΔCEOAE = ∑*_i_ w*(*f_i_*) ΔCEOAE*_i_*,where the sum is taken over all the retained bins.

**Figure 4:**
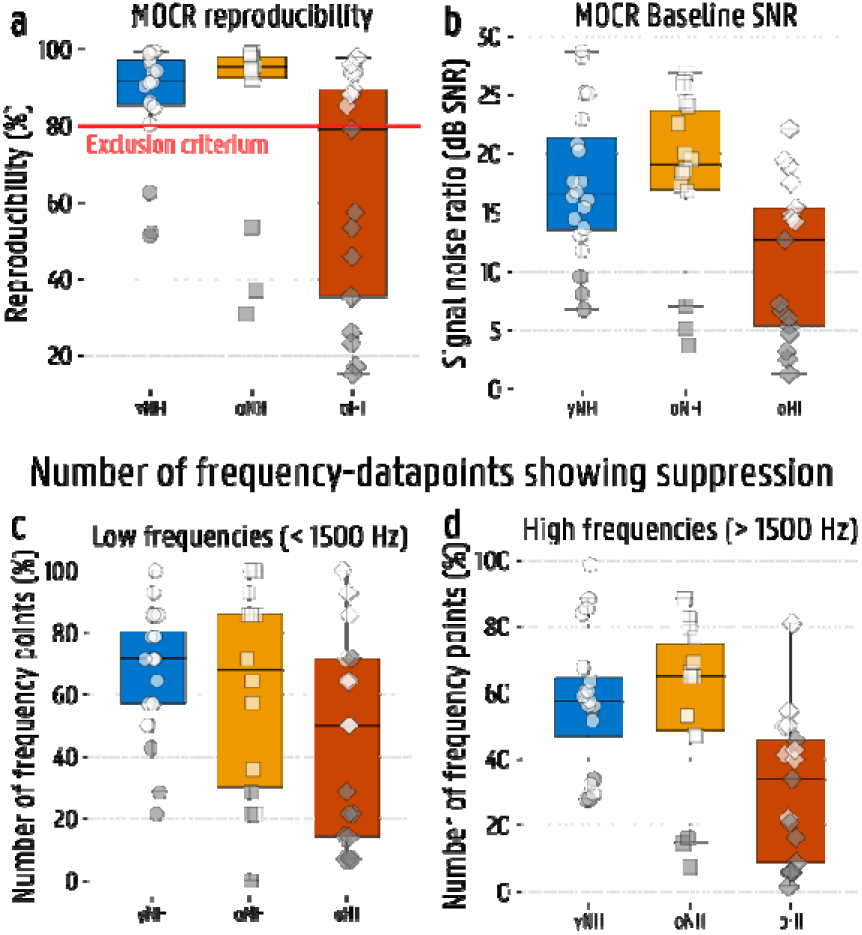
Visualisation of included and excluded medial olivocochlear reflex (MOCR) measurements. (a) Boxplots of MOCR reproducibility of the two buffers, with 80% as minimal inclusion criterium. Excluded measurements are indicated with grey scatters in all four plots. (b) Boxplots of the signal-to-noise ratio (SNR) of the MOCR baseline measurement. (c, d) Boxplots of the number of frequency-datapoints between 600 and 6000 Hz left for each subject after excluding frequencies with SNR < 12 dB at baseline. (c) At low frequencies below 1500 Hz (14 in total). (d) At high frequencies above 1500 Hz (68 in total). (yNH = young normal hearing (N=20), oNH = older normal hearing (N=14), oHI = older hearing impaired (N=17)

Figure 4c and d represent the number of datapoints left after excluding the frequencies with a spectral baseline SNR < 12 dB. MOCR-induced suppression (ΔCEOAE) was the weighted average of the per-bin relative changes. Low-and high-frequency ΔCEOAE values were obtained analogously using frequency-restricted, re-normalized weights.

### MEMR contamination within MOCR measurement

To assess the extent to which potential MEMR activity affected the MOCR measurements, the MOCR was measured using two noise levels in a selection of normal-hearing subjects: 60 dB SPL, where MEMR activation is less expected, and 75 dB SPL, where MEMR activation is more likely. To examine the impact of MEMR on the stimulus, the 0-2 ms time window of the recording was analysed, which corresponds to the window where the evoking click was present. Within this window, the root mean square (RMS) of the difference, baseline minus elicited response, was calculated and used as a measure of possible MEMR co-activation within of the MOCR measurement. A similar technique was earlier described in MOCR-related studies (Abdala et al., 2014; Abdala, Mishra, & Garinis, 2013; Boothalingam et al., 2019). The 75-dB measurements were performed on 14 yNH and 6 oNH subjects.

### Statistical analysis

All statistical analyses were performed in Python. Prior to group comparisons, the distribution of each variable was assessed using the Shapiro-Wilk test for normality (*p* > .05) and Levene’s test for homogeneity of variances *(p* > .01). When assumptions of normality or homogeneity of variance were violated, a Box–Cox transformation was applied.

To assess age-and audiometry-related effects (yNH vs. oNH; oNH vs. oHI), one-way ANOVAs were performed for each variable. When a significant main effect of group was observed, planned pairwise comparisons were performed for the contrasts yNH vs. oNH and oNH vs. oHI using independent-samples Student’s *t* tests with Holm correction for multiple comparisons. Effect sizes for ANOVA results were reported as partial eta squared (η*²p*). Effect sizes for planned pairwise contrasts were reported as Hedges’ *g*.

For MEMR and MOCR variables containing repeated measurements across elicitor side (ipsilateral, contralateral, and/or bilateral conditions), mixed-design ANOVAs were performed with group (yNH, oNH, oHI) as a between-subjects factor and side as a within-subjects factor. When significant group effects were observed, planned post-hoc comparisons between groups (yNH vs. oNH and oNH vs. oHI) were performed using independent-samples Student’s *t* tests with Holm correction. Effect sizes were reported as partial eta squared (η²p) for ANOVA effects and Hedges’ *g* for pairwise comparisons. Statistical significance was set at *p* < .05.

To assess whether age- and audiometry-related group differences persisted after accounting for other aspects of auditory function, a hierarchical covariate-adjustment analysis was performed for each MEMR and MOCR contrast. Covariates were added sequentially in order of increasing physiological non-specificity: cochlear outer hair cell function (mean DPOAE), auditory nerve/brainstem synchrony (EFR amplitude at 4000 Hz), and audiometric sensitivity (HF audiometry, EHF audiometry), reflecting a progression from cochlear measures to more full-pathway auditory detection. At each step, the adjusted group difference, effect size (Hedges’ g), and Bonferroni-corrected p-value (three comparisons) were recorded to track attenuation or persistence of the contrast. To test whether each covariate significantly improved model fit, nested model comparisons (F-tests) were performed at each step, analogous to R²-change testing in hierarchical regression. Multicollinearity at each step was assessed using the variance inflation factor (VIF), with values ≥ 5 flagged as indicating collinearity that may destabilize effect-size estimates.

For the two-group comparisons between MOCR with 60- and 75-dB elicitor levels, a student’s dependent-samples *t*-test was used with α = .05. Correlations were examined using Pearson’s or Spearman’s correlation coefficients based on normality.

## Results

### Hearing status

The audiometric and sensorineural (DPOAE and EFR) hearing status of the three subject groups are described in Figure 5, 6 and 7 respectively and summarized in Table 2. Audiometric thresholds and DPOAE amplitudes were divided between low-(LF, ≤ 1500 Hz) and high-frequency (HF, > 1500 Hz) means. Significant group-effects were observed for all these markers; LF audiometry (*F*(2, 48) = 18.16, *p* < .001, η*²p* = 0.431), HF audiometry (*F*(2, 48) = 49.95, *p* < .001, η*²p* = 0.675), EHF audiometry (*F*(2, 48) = 76.45, *p* < .001, η*²p* = 0.761), LF DPOAE (*F*(2, 48) = 6.37, *p* = .004, η*²p* = 0.210), HF DPOAE (*F*(2, 48) = 24.20, *p* < .001, η*²p* = 0.502) and EFR (*F*(2, 46) = 13.82, *p* < .001, η*²p* = 0.375).

**Figure 5:**
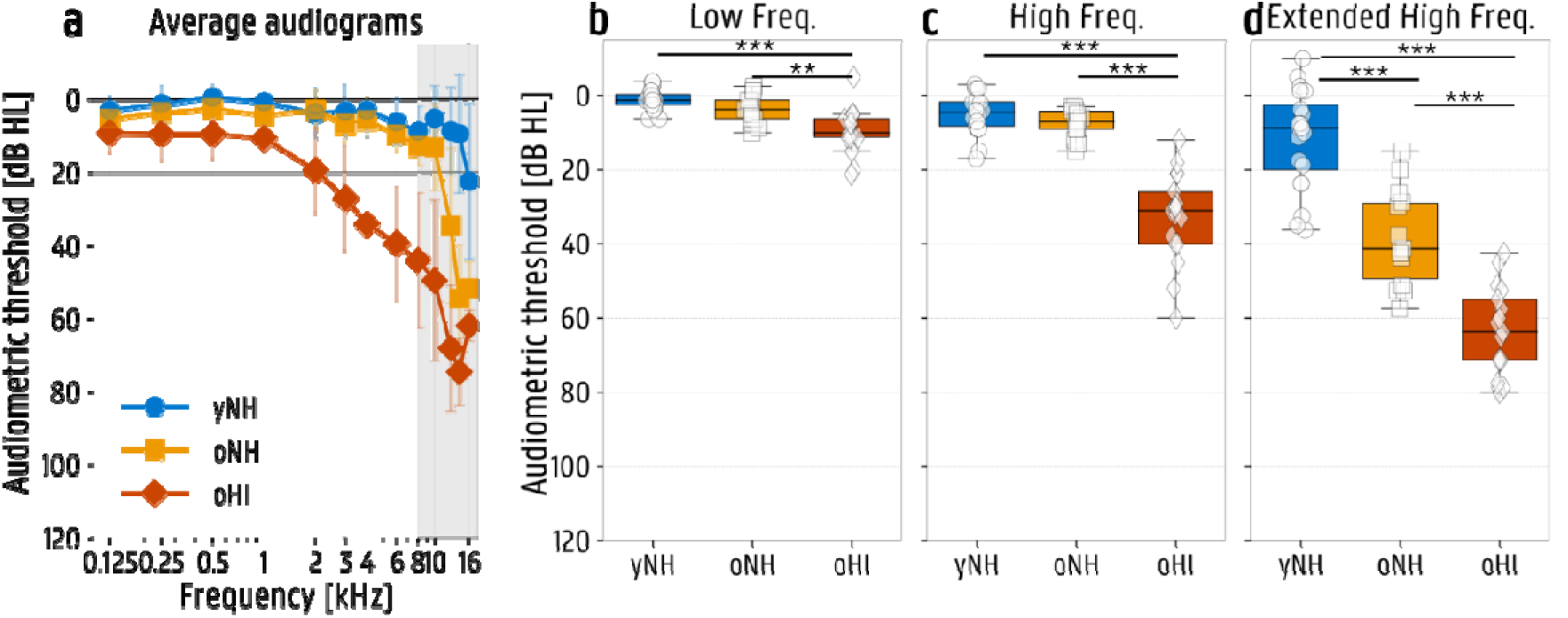
Tonal audiometric thresholds for the three test groups; yNH = young normal hearing (N=20), oNH = older normal hearing (N=14), oHI = older hearing impaired (N=17). (a) Average audiometric thresholds per frequency, per group. (b) Boxplots of mean thresholds at low frequencies between 0.125 and 1 kHz. (c) Boxplots of mean thresholds at high frequencies between 2 and 8 kHz. (d) Boxplots of mean thresholds at extended high frequencies between 10 and 16 kHz. *p < .05; **p < .01; ***p < .001.

**Figure 6:**
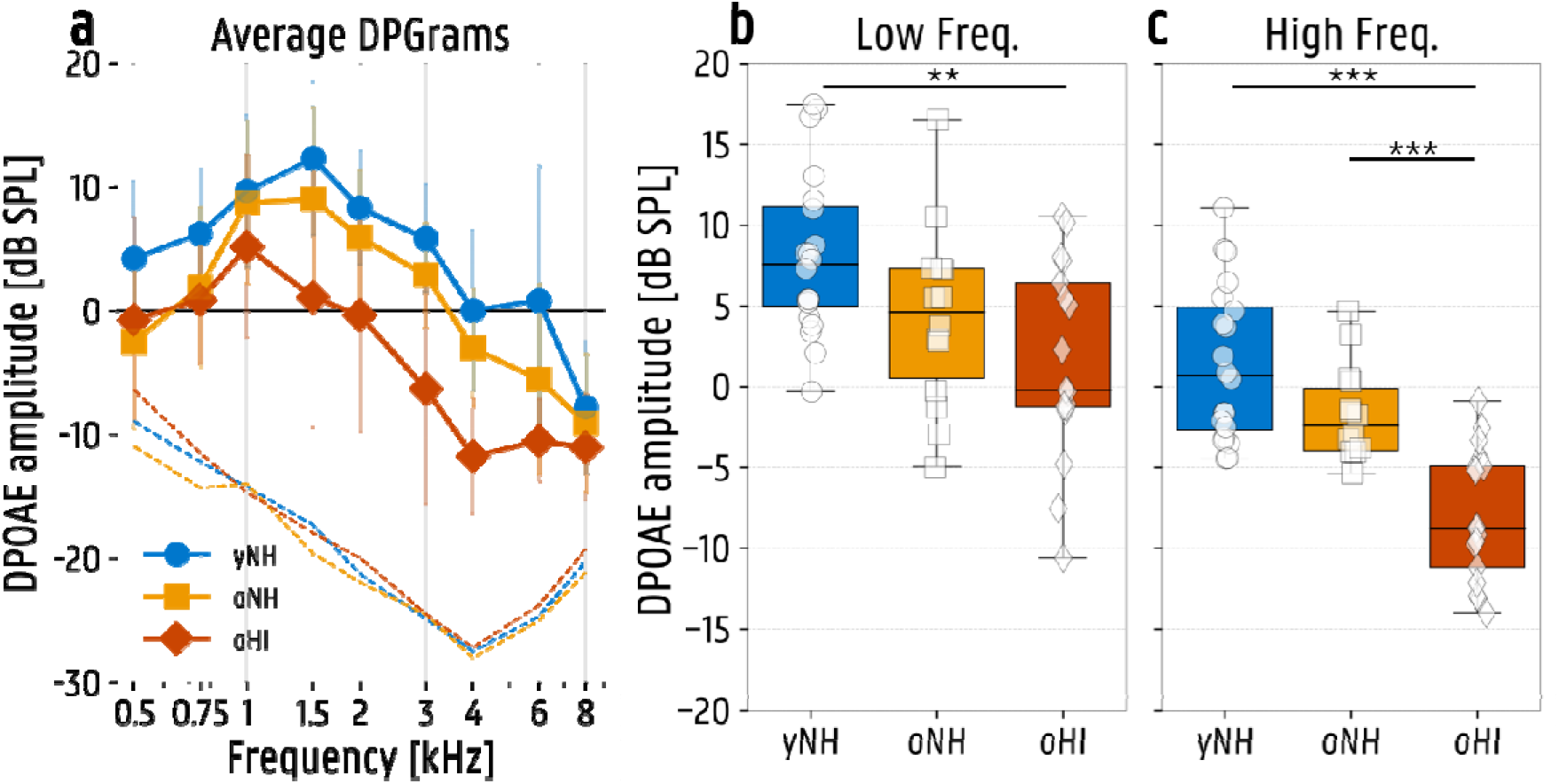
Distortion product otoacoustic emissions (DPOAE) for the three test groups yNH = young normal hearing (N=20), oNH = older normal hearing (N=14), oHI = older hearing impaired (N=17). (a) Average DPOAE amplitudes per frequency, per group with the noise-floors shown as dashed lines. (b) Boxplots of mean DPOAE amplitudes at low frequencies between 0.5 and 1.5 kHz. (c) Boxplots of mean DPOAE amplitudes at high frequencies between 2 and 8 kHz. *p < .05; **p < .01; ***p < .001

**Figure 7:**
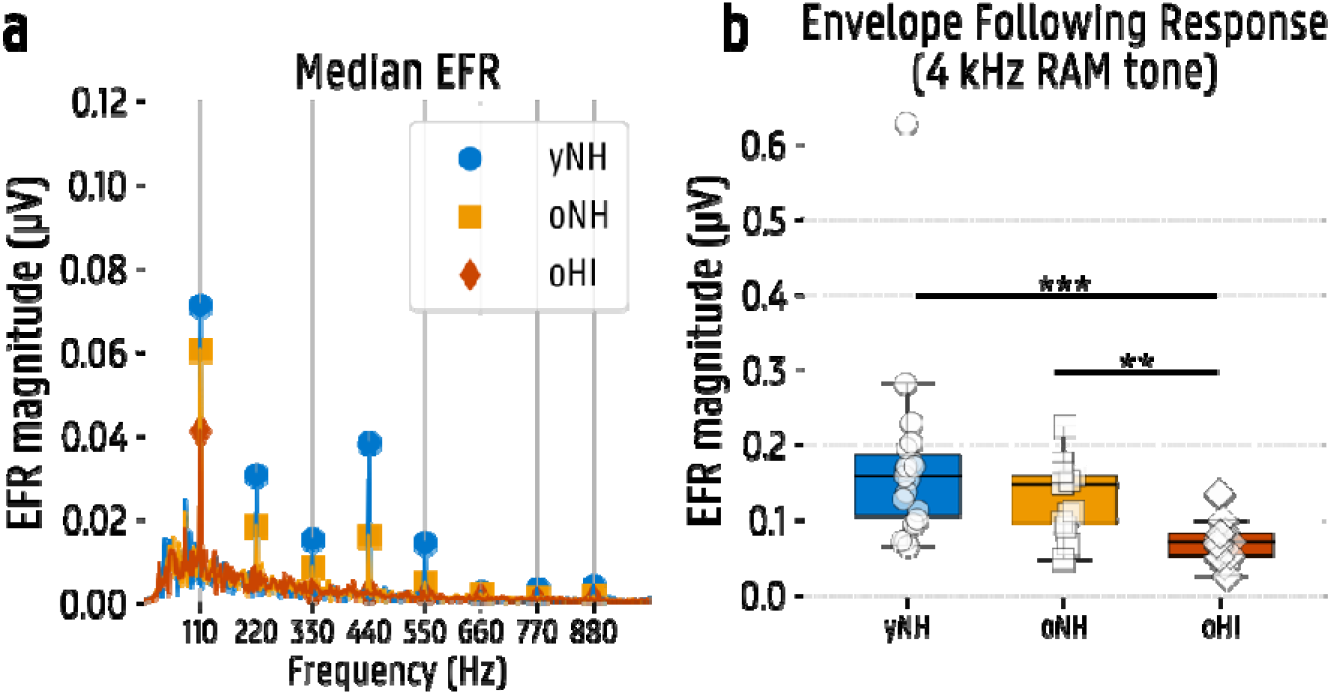
(a) Median spectral EEG-response to the envelope following response (EFR) stimulus. Harmonics of the modulation frequency of 110 Hz are indicated with markers. (b) Boxplots of the EFR magnitude for the three test groups, which is the sum of the first four harmonics. yNH = young normal hearing (N=19), oNH = older normal hearing (N=13), oHI = older hearing impaired (N=17). RAM = rectangular amplitude modulated. *p < .05; **p < .01; ***p < .001.

**Table 2:** Summary of post-hoc age- and audiometry-related group effects on audiometry, DPOAE and EFR.

|  | Age-related effect<br>yNH vs oNH | Audiometry-related effect<br>oNH vs oHI | yNH vs oHI |
| --- | --- | --- | --- |
| *LF audiometry* | NO<br>$t(23) = -2.41, p = .073, g = -0.86$ | YES<br>$t(27) = -3.26, p = .009, g = -1.10$ | YES<br>$t(22) = -5.39, p < .001, g = -1.83$ |
| *HF audiometry* | NO<br>$t(29) = -2.11, p = .132, g = -0.64$ | YES<br>$t(29) = -9.51, p < .001, g = -3.24$ | YES<br>$t(33) = -9.02, p < .001, g = -2.82$ |
| *EHF audiometry* | YES<br>$t(29) = -5.91, p < .001, g = -2.00$ | YES<br>$t(27) = -5.55, p < .001, g = -1.97$ | YES<br>$t(35) = -12.51, p < .001, g = -4.00$ |
| *LF DPOAE* | NO<br>$t(26) = 2.03, p = .158, g = 0.71$ | NO<br>$t(29) = 1.27, p = .641, g = 0.44$ | YES<br>$t(31) = 3.50, p = .004, g = 1.15$ |
| *HF DPOAE* | NO<br>$t(32) = 2.38, p = .071, g = 0.75$ | YES<br>$t(29) = 4.88, p < .001, g = 1.67$ | YES<br>$t(35) = 6.47, p < .001, g = 2.06$ |
| *EFR 4000 Hz* | NO<br>$t(29) = 1.24, p = .673, g = 0.42$ | YES<br>$t(26) = 3.70, p = .003, g = 1.32$ | YES<br>$t(34) = 5.05, p < .001, g = 1.64$ |

MEMR = middle ear muscle reflex; MOCR = medial olivocochlear reflex; LF = low frequencies; HF = high frequencies; DPOAE = distortion product otoacoustic emissions, EFR = envelope following response, EHF = extended high frequencies.

Post-hoc age-related (yNH vs. oNH) differences were significant for EHF audiometry only (Table 2). Audiometry-related differences (oNH vs. oHI) were observed for all markers (LF audiometry, HF audiometry, EHF audiometry, HF DPOAE and EFR), except for LF DPOAE. All markers showed a significant difference between yNH and oHI.

### Reflex measurements

#### Effect of age and hearing loss on MEMR and MOCR

To address the question whether MEMR and MOCR measurements are related to age-related subclinical and audiometric clinical hearing loss, we investigated the age-related difference between the yNH and oNH groups and audiometry-related differences between oNH and oHI groups.

For MEMR, thresholds and amplitudes were analysed for the ipsilateral and contralateral elicitor condition. We used a 3×2 mixed-design ANOVA, with group (three levels) as the between-subjects factor and elicitor side (ipsilateral vs. contralateral) as the within-subjects factor (Figure 8). The MEMR thresholds showed significant group effects (*F*(2, 36) = 4.90, *p* = .013, η*²p* = 0.214), but no side-related differences or interaction effect (*p* > .05). MEMR amplitudes at 90-dB elicitor level showed significant differences for both group (*F*(2, 36) = 3.68, *p* = .035, η*²p* = 0.170) and side effects (*F*(1, 36) = 15.06, *p* < .001, η*²p* = 0.295). Again, no interaction effects were observed, suggesting that group effects were consistent across elicitor conditions (*p* > .05). Post-hoc analysis showed an age-related significant difference in MEMR amplitude, but not in MEMR threshold (Table 3). No significant audiometry-related post-hoc differences were present in either MEMR threshold or amplitude (*p* > .05).

**Figure 8:**
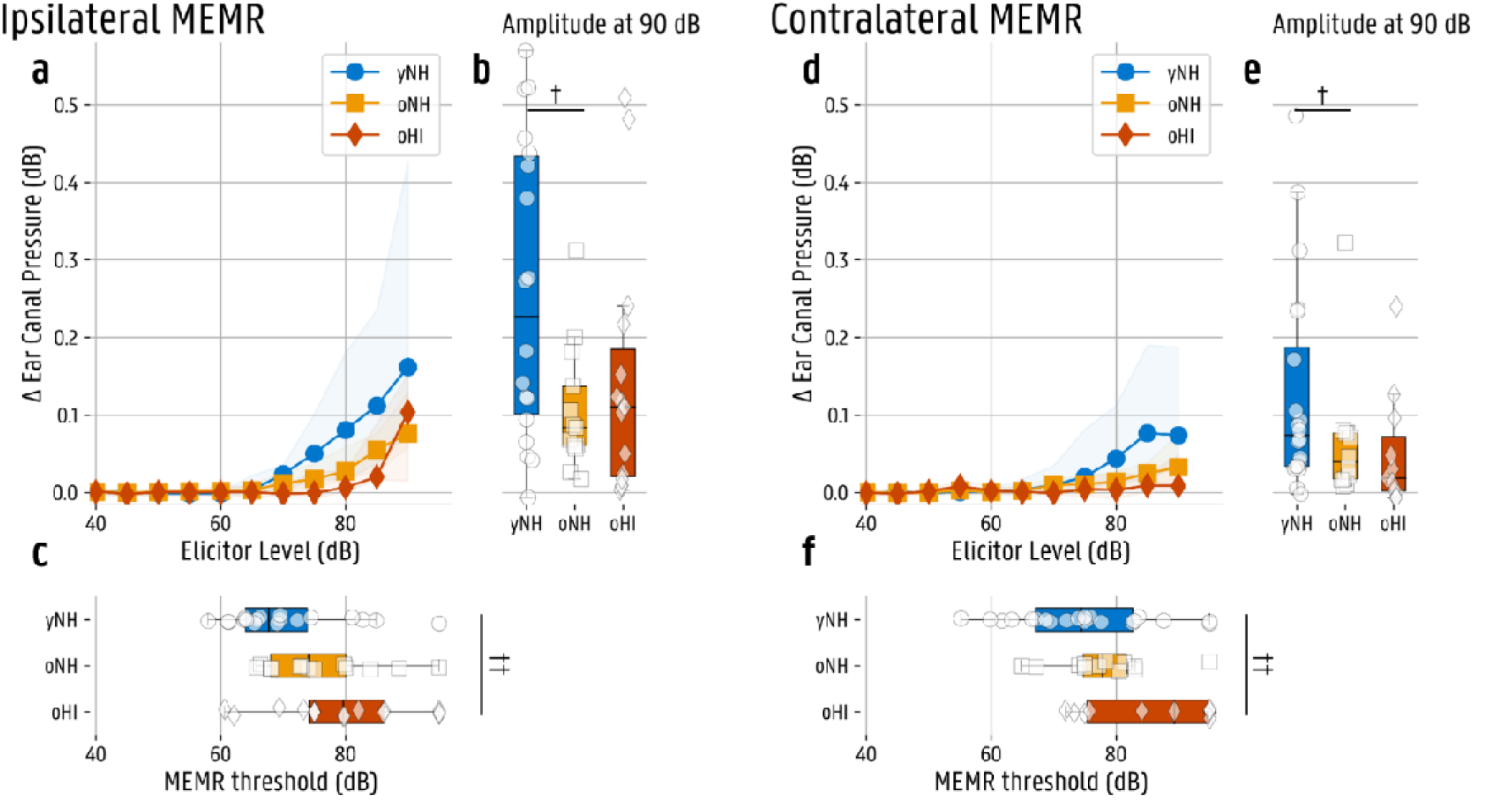
Ipsilateral and contralateral middle ear muscle reflex (MEMR) outcome of the three test groups; yNH = young normal hearing (ipsi N=18, contra N=20), oNH = older normal hearing (ipsi N=13, contra N=12), oHI = older hearing impaired (ipsi N=15, contra N=11). (a) Median ipsilateral MEMR growth function with interquartile range (IQR) shading. (b) Boxplots of ipsilateral ear-canal pressure change at 90 dB SPL elicitor level. (c) Boxplots of ipsilateral MEMR thresholds. (d) Median contralateral MEMR growth function with IQR shading. (e) Boxplots of contralateral ear-canal pressure change at 90 dB SPL elicitor level. (f) Boxplots of contralateral MEMR thresholds. ANOVA group-effects using average of ipsilateral and contralateral data, ^†^p < .05; ^††^p < .01; ^†††^p < .001.

**Table 3:** Summary of age- and audiometry-related group effects on MEMR and MOCR following sequential adjustment for sensorineural covariates.

|  | Age-related effect<br>yNH vs oNH | Audiometry-related effect<br>oNH vs oHI | yNH vs oHI |
| --- | --- | --- | --- |
| **MEMR threshold** | NO<br>$t(60) = -2.07, p = .128, g = -0.50$ | NO<br>$t(47) = -1.76, p = .255, g = -0.48$ | YES<br>$t(55) = -3.55, p = .002, g = -0.88$ |
| **MEMR amplitude** | YES, but<br>$t(60) = 2.57, p = .038, g = 0.61$<br>Significance disappeared after DPOAE correction. | NO<br>$t(46) = 0.11, p = 1.000, g = 0.03$ | NO<br>$t(54) = 2.31, p = .075, g = 0.58$ |
| **MOCR LF** | NO<br>$t(82) = -0.63, p = 1.000, g = -0.13$ | YES<br>$t(45) = 2.94, p = .015, g = 0.80$<br>Significance disappeared after DPOAE correction. Hedges' $g$ remained > 0.5 until correction for HF audiometry, suggesting a potentially underpowered comparison. | NO<br>$t(61) = 2.08, p = .126, g = 0.45$ |
| **MOCR HF** | YES, but<br>$t(80) = 2.87, p = .016, g = 0.60$<br>Significance disappears after correction for HF audiometry. | NO<br>$t(37) = 1.37, p = .536, g = 0.39$ | YES<br>$t(40) = 3.36, p = .005, g = 0.87$ |

MEMR = middle ear muscle reflex; MOCR = medial olivocochlear reflex; LF = low frequencies; HF = high frequencies; DPOAE = distortion product otoacoustic emissions, EFR = envelope following response, EHF = extended high frequencies.

For the MOCR, we investigated the ΔCEOAE between the three test groups for the three elicitor conditions used (ipsilateral, contralateral and bilateral), using a 3×3 mixed-design ANOVA, performed for the low-frequency datapoints (< 1500 Hz) and the high-frequency datapoints (> 1500 Hz) separately (Figure 9). The LF MOCR showed no significant group effect (*F*(2, 33) = 1.56, *p* = .226, η*²p* = 0.086), but had a significant side effect (*F*(2, 66) = 40.25, *p* < .001, η*²p* = 0.549) and no interaction effect (*p* > .05). HF MOCR revealed significant group (*F*(2, 33) = 5.19, *p* = .011, η*²p* = 0.239) and side effects (*F*(2, 66) = 28.82, *p* < .001, η*²p* = 0.466), without interaction (*p* > .05). Post-hoc analysis revealed an age-related difference for HF MOCR, and an audiometry-related differences for LF MOCR (Table 3).

**Figure 9:**
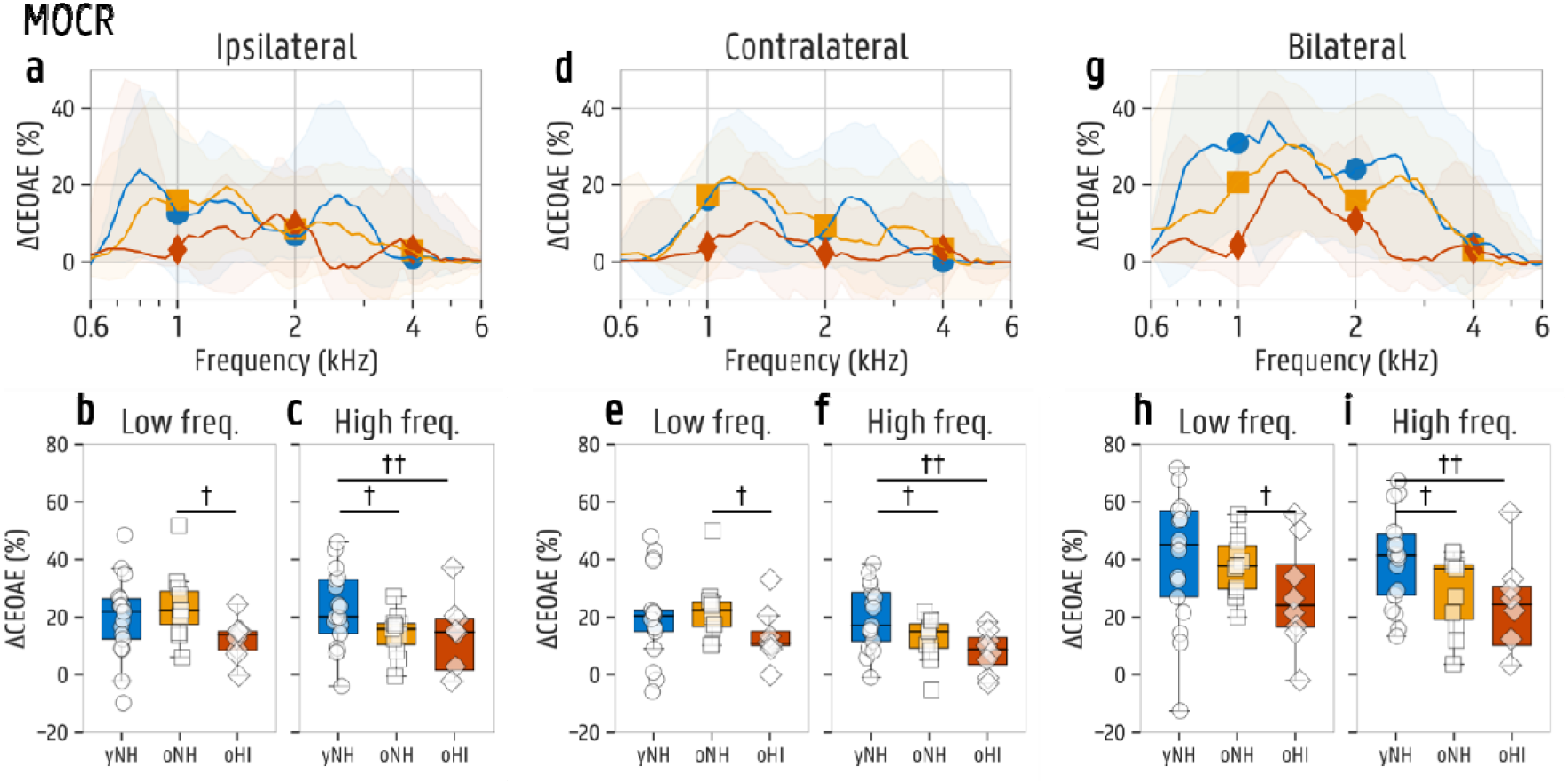
Outcome of the medial olivocochlear reflex (MOCR) with ipsilateral, contralateral and bilateral elicitors, comparing the three test groups; yNH = young normal hearing (N=17), oNH = older normal hearing (N=11), oHI = older hearing impaired (N=8). (a, d, g) Median unweighted relative ipsilateral, contralateral and bilateral MOCR-evoked change in click evoked otoacoustic emissions (CEOAE) per frequency with interquartile range (IQR) shading. (b, e, h) Boxplots of mean weighted relative ipsilateral, contralateral and bilateral MOCR-evoked changes in CEOAE at low frequencies between 0.6 and 1.5 kHz. (c, f, i) Boxplots of mean ipsilateral, contralateral and bilateral MOCR-evoked changes in CEOAE at high frequencies between 1.5 and 6 kHz. ^†^p < .05; ^††^p < .01; ^†††^p < .001.

#### Hierarchical adjustment for afferent sensorineural markers

Similar group comparisons were performed on MEMR and MOCR, while adjusting for afferent sensorineural and audiometric covariates in a hierarchical way (see details in Statistical analysis section). These analyses are visualized in Figure 10 and summarized in Table 3.

**Figure 10:**
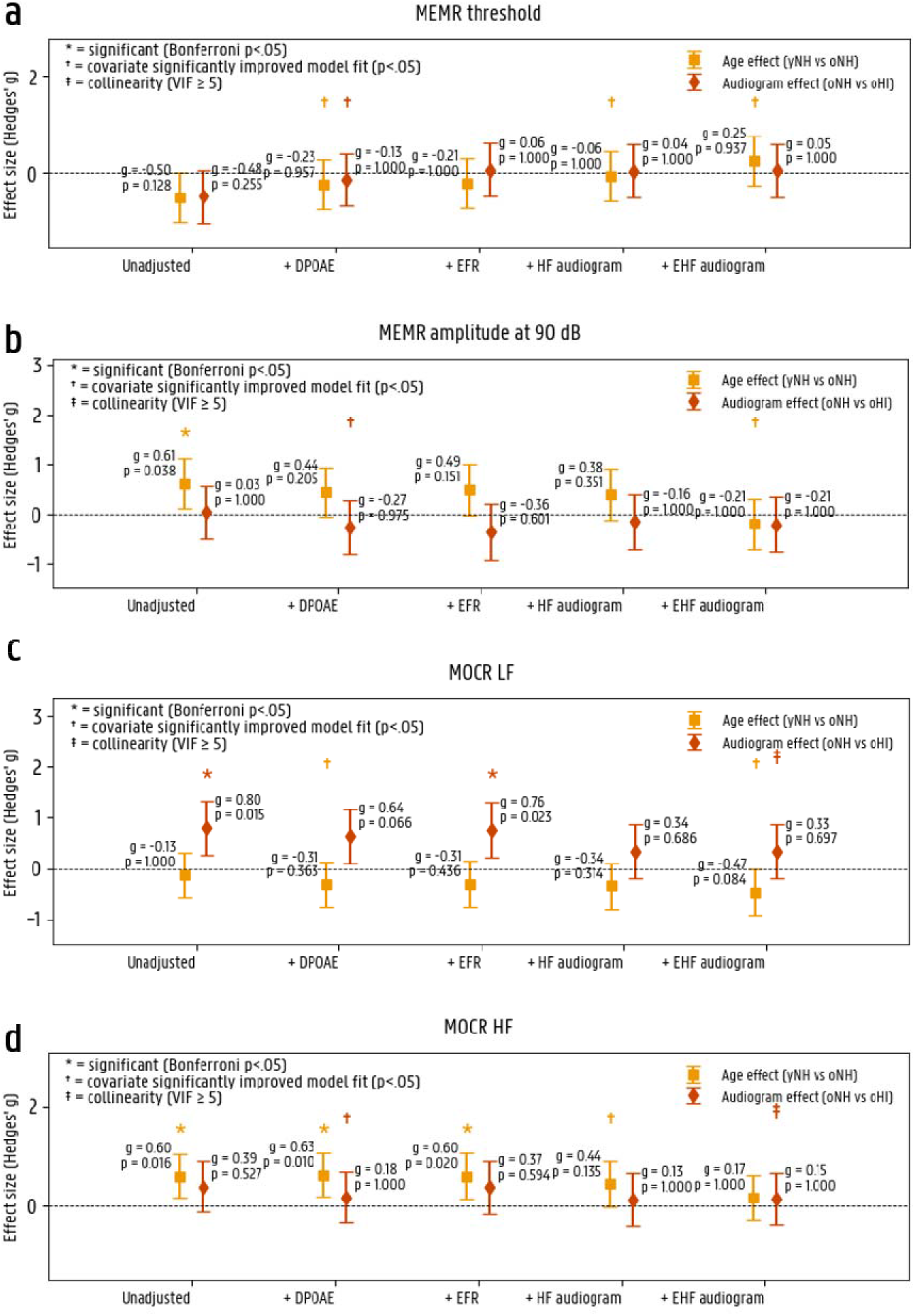
Stepwise evaluation of group differences in auditory reflex measures after hierarchical covariate adjustment; middle ear muscle reflex (MEMR) threshold, MEMR amplitude, medial olivocochlear reflex (MOCR) low-frequent (LF) and MOCR high-frequent (HF). Effect sizes (Cohen’s d) are shown for the contrast between younger normal-hearing (yNH) and older normal-hearing (oNH) participants (age-related effect) and between oNH and older hearing-impaired (oHI) participants (audiometry-related effect). Error bars represent 95% confidence intervals. Models were hierarchically adjusted for sensorineural auditory measures (distortion product otoacoustic emissions (DPOAE);envelope following responses (EFR)) and mean audiometric thresholds (high-frequency (HF); extended high-frequency (EHF)). Asterisks indicate significant adjusted group effects after Bonferroni correction for multiple comparisons (three; as in the original ANOVA analysis). Daggers indicate that inclusion of the covariate significantly improved model fit based on nested F-tests. Double daggers indicate potential multicollinearity (variance inflation factor ≥5). Positive Cohen’s d values indicate higher values in the first group of each contrast (yNH > oNH or oNH > oHI).

For MEMR thresholds, neither the age-related contrast nor the audiometry-related contrast reached significance in the unadjusted analysis (*p* > .05). Both effects moved further from significance following adjustment for DPOAE, and remained non-significant through the remaining covariate steps (Figure 10a). Across all steps, effect sizes stayed small as well (*g* < 0.5).

The MEMR amplitude at 90 dB showed a significant age-related effect in the unadjusted analysis (*g* = 0.61, *p* = .038), and lost significance after DPOAE correction (*p* > .05) (Figure 10b). While adding EFR and HF audiometry did not significantly improve the model, EHF audiometry did (Δ*F* = 11.61, Δ*p* = .001). The audiometry-related contrast was non-significant in the unadjusted model (*g* = -0.02, *p* = 1.000), and remained non-significant throughout all adjustment steps (*p* > .05).

MOCR in the low frequencies (Figure 10c) showed a significant audiometry-related contrast in the unadjusted analysis (*g* = 0.80, *p* = .015), and lost significance after DPOAE correction (*g* = 0.64, *p* > .05). Given that effect size remained moderate-to-large (*g* > 0.5), the present sample may have been underpowered to detect significance at this point. The age-related contrast was non-significant in the unadjusted model (*g* = -0.13, *p* = 1.000), and remained non-significant following adjustment for DPOAE, EFR and HF audiograms (*p* > .05).

For MOCR in the high frequencies (Figure 10d), the age-related contrast was significant in the unadjusted analysis (*g* = 0.60, *p* = .016), and remained significant following DPOAE (*g* = 0.63, *p* = .010) and EFR adjustment (*g* = 0.60, *p* = .020), but became non-significant after adjustment for HF audiogram (*p* > .05; Δ*F* = 7.51, Δ*p* = .008) indicating that HF audiometric thresholds account for a substantial portion of this age-related effect. The audiometry-related contrast was non-significant in the unadjusted analysis (*p* > .05), and remained non-significant across all adjustment steps.

#### MEMR co-activation within MOCR measurement

To study the potential MEMR contamination during an MOCR measurement, we performed an MOCR measurement at two elicitor levels: 60 and 75 dB SPL. At 60 dB SPL, MEMR activation was considered unlikely, whereas at 75 dB SPL, MEMR activation was expected in most subjects. MEMR and MOCR responses at 60 dB and 75 dB are visualized in Figure 11. Within the yNH group, 1 subject (6.7%) had an MEMR amplitude above the threshold criteria of 0.02 dB at 60 dB and 10 subjects had a present MEMR at 75 dB (66.7%; Figure 11 c, d). Within the oNH group, no subjects had a present MEMR at 60 dB and 3 subjects at 75 dB (50%; Figure 11 c, d). This means that we expect co-activation in one subject at 60 dB and in around half of the subjects at 75 dB. Performing the MOCR with a 75-dB elicitor noise resulted in significantly stronger low-frequency suppression (*M* = 41.81, *SD* = 24.66) compared to the 60-dB condition (*M* = 25.51, *SD* = 12.51; *t(*15) = -2.93, *p* = .010, *d* = -0.73; Figure 11 e, f). Also for the high frequencies, a similar significant increase in suppression was observed between 60 dB (*M* = 19.39, *SD* = 12.47) and 75 dB (*M* = 30.62, *SD* = 18.24; *t(*15) = -3.02, *p* = .009, *d* = -0.75; Figure 11 e, g). MEMR co-activation was thus visible in both high and low-frequency analysis of the MOCR, and most prominent for the low frequencies.

**Figure 11:**
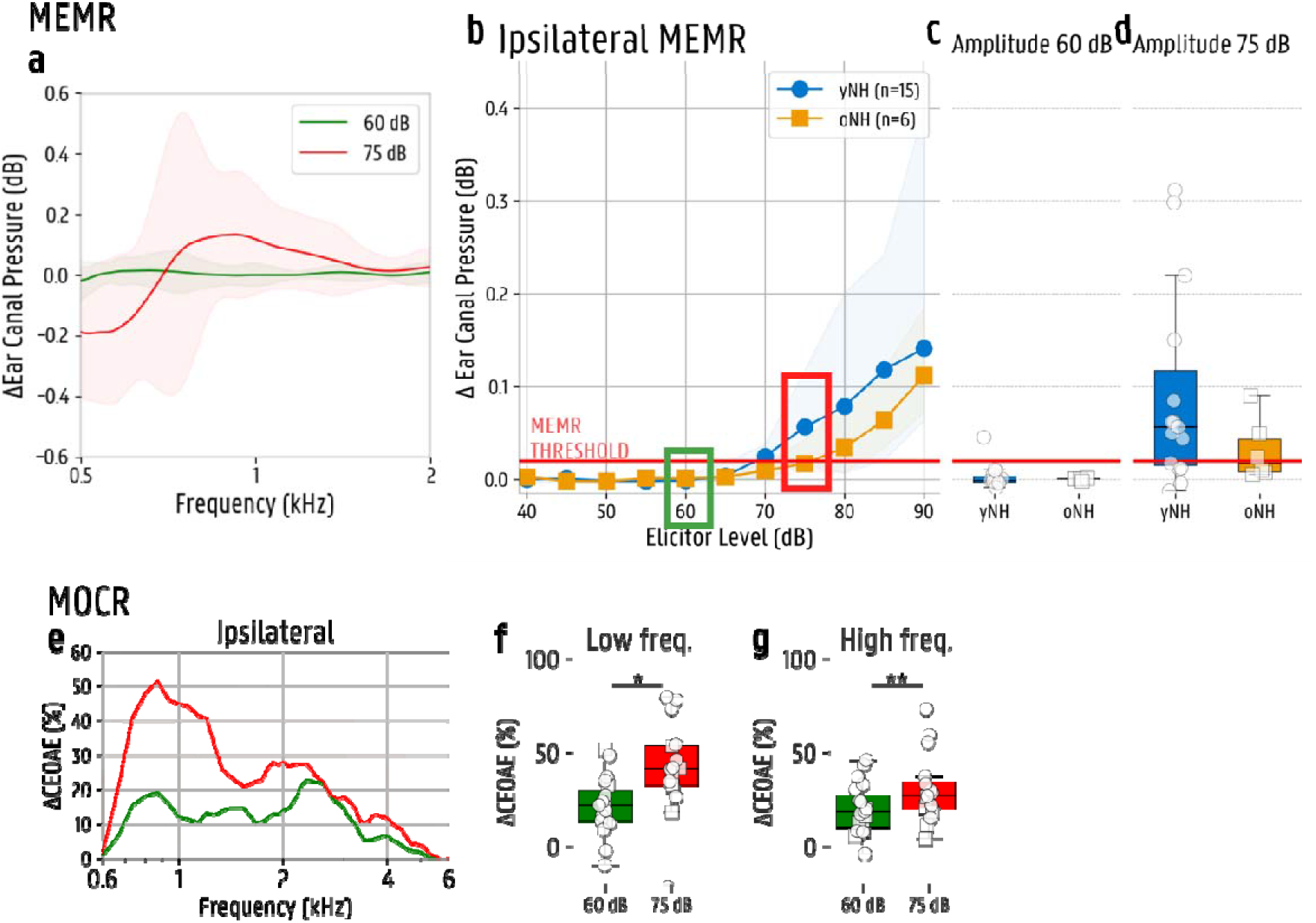
Visualization of the middle ear muscle reflex (MEMR) and medial olivocochlear reflex (MOCR) response at 60 dB SPL and 75 dB SPL elicitor level. (a) MEMR difference in ear-canal pressure (baseline minus evoked response) per frequency. 60 dB is presented in green and shows no reflex, while 75 dB in red shows a frequency change due to the MEMR contraction. (b) MEMR growth function with indication of 60- and 75-dB elicitor levels (green and red box) and the threshold criteria of 0.02 dB (red line). yNH = young normal hearing; oNH = older normal hearing. (c, d) Boxplots of the MEMR amplitude at an elicitor level of 60 dB and 75 dB. (e) Relative MOCR-induced suppression for the 60 dB (green) and 75 dB (red) condition in the frequency domain. (f, g) Boxplots of the MOCR-induced suppression comparing 60 dB and 75 dB elicitor levels for low- and high-frequency responses separately. yNH = young normal hearing (N=14), oNH = older normal hearing (N=9). *p < .05; **p < .01; ***p < .001.

With the reasoning in mind that a contraction of the MEMR would directly reduce the click response in the ear canal, it is expected that MEMR co-activation would be visible within the first 2 ms of the click response (Figure 12 a, b). To check this assumption, the relative click-response difference (baseline minus elicited response), from 0 to 2 ms, was used as a possible marker for MEMR co-activation (see blue difference-line in the click domain in Figure 12 a and b). This marker was significantly higher at 75 dB elicitor level (*M* = 0.051, *SD* = 0.0871) compared to 60 dB (*M* = 0.003, *SD* = 0.0110; *t(*15) = -2.37, *p* = .031, *d* = -0.59; Figure 12c). At 75 dB, this marker correlated strongly with the separately measured MEMR amplitude at 75-dB elicitor level (*r*(14) = .92, *p* < .001; Figure 12e), reflecting co-activation of the MEMR reflex within the MOCR measurement at 75-dB elicitor noise. The MOCR click difference at 60-dB elicitor level also correlated moderately with the MEMR response at 60dB or 75 dB (*r*(17) = .61, *p* = .005; Figure 12d). This aligns with the hypothesis that MEMR contamination reduces the effective click level used in an MOCR measurement, and that this reduction becomes stronger with higher MEMR contraction based on the MEMR amplitude.

**Figure 12:**
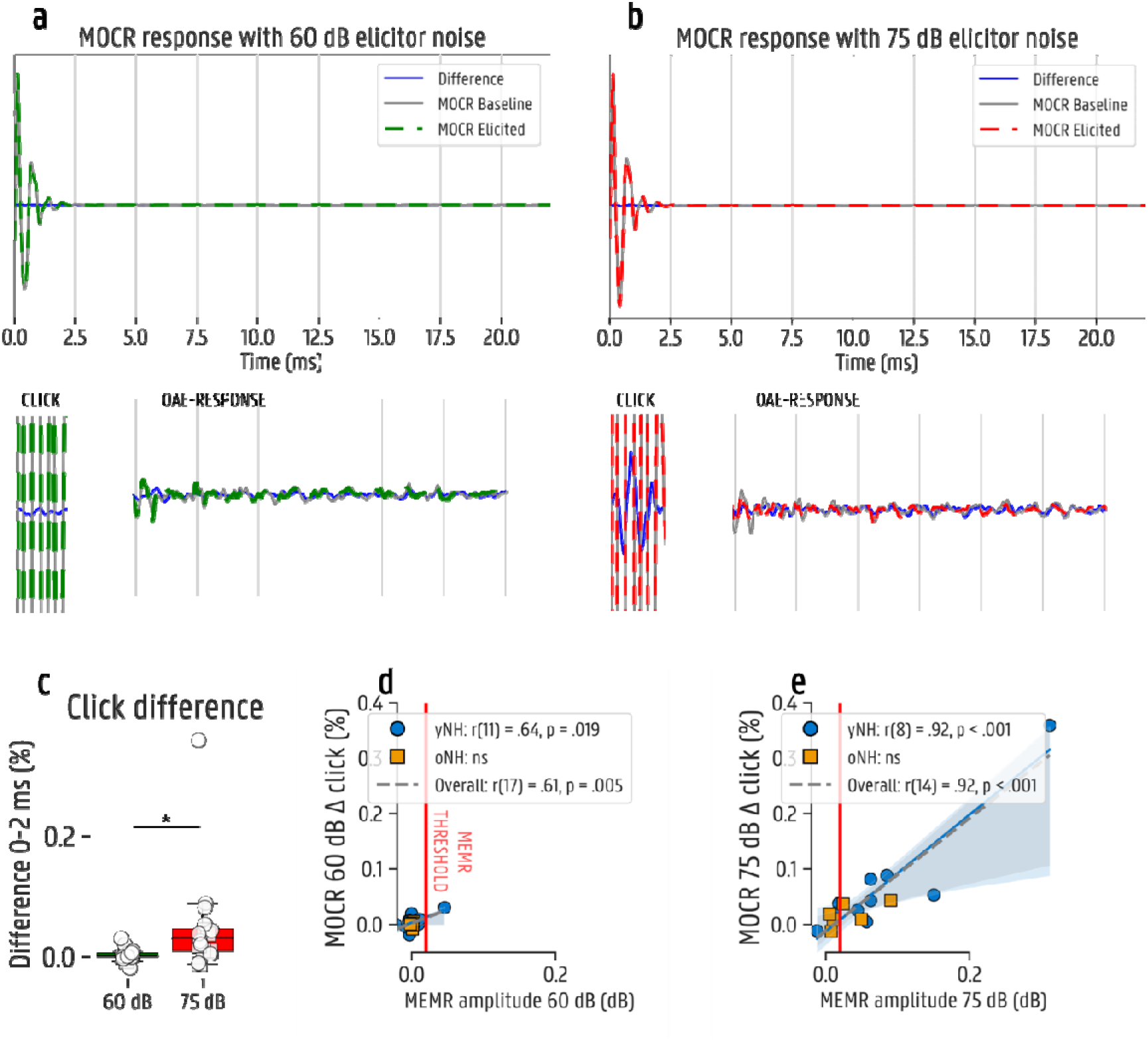
(a, b) Visualization of the medial olivocochlear reflex (MOCR) click response for baseline, elicited and the difference, baseline minus elicited condition, at 60 dB and 75 dB. The click domain is indicated from 0 to 2 ms, while the CEOAE response is calculated from 5 to 20 ms. (c) Boxplots of the mean difference, baseline minus elicited condition, from 0 to 2 ms at 60- and 75-dB elicitor level. (d, e) Scatter dot correlation plots between the MOCR click difference and MEMR amplitude at 60- and 75-dB elicitor level. yNH = young normal hearing; oNH = older normal hearing. yNH = young normal hearing (N=14), oNH = older normal hearing (N=9). *p < .05; **p < .01; ***p < .001.

## Discussion

In this study, we explored how MEMR and MOCR measurements are affected by age and audiometric hearing loss. We compared three test groups (yNH, oNH, and oHI) and investigated whether the observed age-and audiometry-related effects survived after correction for DPOAE, EFR, HF audiometry, and EHF audiometry. To improve measurement validity of these reflex measurements, we explored a potential marker for MEMR co-activation during MOCR measurements to improve measurement validity.

### MEMR as a potential marker for age-related subclinical hearing damage

While MEMR thresholds were not able to detect significant group differences, MEMR amplitudes showed a significant age-related group difference. The other two markers for sensorineural damage (DPOAE and EFR) did not reveal a significant age effect in this dataset, while EHF audiometry did. The age-related difference in MEMR amplitude was significantly reduced by correcting for DPOAE and EHF audiometry. This suggests that the MEMR amplitude could be related to subtle cochlear damage.

Previous studies hypothesized that MEMR may serve as a marker for cochlear synaptopathy. Animal studies have shown that MEMR strength is reduced after noise-exposure or aging, sometimes providing a better predictor than the ABR wave I amplitude (Bharadwaj et al., 2022; Bharadwaj et al., 2019; Kobler et al., 1992; Valero, Hancock, & Liberman, 2016; Valero et al., 2018). The underlying logic is that the high-threshold, low spontaneous rate fibers, which are believed to be the first to degenerate as a result of noise exposure or aging (Furman, Kujawa, & Liberman, 2013; Reijntjes et al., 2025), may also be important aspects of the MEMR pathway (Kobler et al., 1992; Valero, Hancock, & Liberman, 2016; Valero et al., 2018). Nevertheless, the marker that we used to address cochlear synaptopathy (EFR), did not significantly explain the MEMR-related age difference. Similarly, Bramhall et al. (2022) did not find a correlation between the MEMR and EFR strength in veterans.

This absence of correlations within the normal-hearing groups could be due to age ranges, heterogeneity of synaptopathy degrees within the groups, and confounding factors. Furthermore, MEMR and EFR measurements differ as they use different measurement techniques (neural response vs change in ear canal pressure), involve different neural structures and rely on different stimulus probes. The MEMR pathway involves the superior olivary complex and returns via the efferent facial nerve, whereas the EFR also reflects more central brainstem nuclei, such as the inferior colliculus. Thus, MEMR may reflect the effect of cochlear synaptopathy earlier in the afferent pathway but is also influenced by efferent activity and middle-ear reflections, also mentioned by Bramhall et al. (2022). Additionally, MEMR was elicited with broadband noise, whereas the EFR is evoked with a 4-kHz amplitude-modulated tone. Future studies should compare 4-kHz EFR with MEMR elicited by a 4-kHz stimulus to assess region-specific synaptopathy.

In general, a better frequency specificity of the MEMR elicitor would be beneficial to further discover the influence of cochlear damage, since EHF audiometric thresholds seem to be related to the MEMR. Age-related hearing loss primarily affects high-frequency regions, while low-frequency thresholds often remain normal. Consequently, MEMR responses in this study may predominantly reflect low-frequency cochlear regions, potentially reducing sensitivity to high-frequency cochlear damage. Future studies should use frequency-specific MEMR elicitors to determine whether high-frequency MEMR is even better related to markers of sensorineural damage, including outer-hair cell dysfunction, synaptopathy and elevated audiometric thresholds.

### MOCR measurements are not affected by subclinical hearing loss but require adjusted techniques for subjects with clinical hearing loss

For MOCR, age-related differences were only observed in the high frequencies, not in the low frequencies. This difference remained after correction for DPOAE and EFR, and disappeared after correcting for high-frequency audiometric thresholds. This suggests that MOCR measurements, when corrected for CEOAE baseline differences, are relatively robust to cochlear variation in normal-hearing subjects. However, many oHI measurements were excluded due to absent baseline OAEs and poor reproducibility, which points out the potential effect of audiometric hearing loss on the MOCR.

This raises the question whether one should increase the intensity of the elicitor stimulus or probe stimulus based on audiometric thresholds. Earlier research showed that it is possible and advantageous to use higher levels for MOCR testing (Guinan Jr et al., 2003). Future studies should investigate whether MOCR elicitor-level growth functions, as employed in MEMR paradigms, reveal differential patterns across age and hearing loss groups that are not captured by single-level measurements. Lewis (2019) observed a general decrease in MOCR strength with growing click intensities, differing across frequencies. Similar measurements should be conducted in participants with sloping high-frequency hearing losses. An important factor to consider when increasing the elicitor level, is the potential co-activation of the MEMR reflex, which may compromise the MOCR, and is mainly expected in the younger groups, due to overall lower MEMR thresholds.

### MEMR co-activation can be measured by quantifying the click-stimulus window in MOCR measurements

Our results show that the MEMR can interfere with MOCR measurements, and that this interference is detectable in the click-stimulus window during MOCR testing. By analysing the first 2 ms of the click response, we identified a clear marker of MEMR activation. At 75 dB SPL, this early click difference was much larger than at 60 dB SPL and closely matched separately measured MEMR amplitudes. At 75 dB SPL, about half of the subjects showed MEMR activation during the MOCR measurement, while at 60 dB SPL only one subject did. This suggests that 60 dB SPL is generally a safe elicitor level with minimal MEMR influence, but individual testing is still needed to exclude potential outliers. Future work should establish objective criteria to identify and exclude MEMR contamination.

## Conclusion and recommendations for clinical use

Middle ear muscle reflex:

- MEMR thresholds showed no significant age-or audiometry-related effects for MEMR threshold at any stage of the adjustment for covariates. Within the constraints of the present sample size, this pattern does not provide evidence that MEMR threshold serves as a marker of age- or hearing-loss-related auditory deficits.
- MEMR amplitude showed an age-related effect that was attenuated after adjustment for DPOAE and extended high-frequency audiometric thresholds. It is possible that the apparent age effect is at least partly related to cochlear damage, though the present design cannot establish whether this reflects a shared underlying mechanism or not.

Medial olivocochlear reflex:

- High-frequency MOCR showed an age-related effect that was reduced following adjustment for HF audiometric thresholds but not following adjustment for DPOAE or EFR, suggesting that MOCR sensitivity to hearing status appears more closely tied to audiometric than subclinical sensorineural status. Given the contrast with the low-frequency MOCR results, future studies should dissociate low- and high-frequency MOCR more clearly by varying the elicitor.
- MEMR co-activation can be detected in the first ∼2 ms after click presentation and can artificially alter MOCR-induced suppression. Measuring MEMR separately alongside MOCR testing is recommended to check for potential MEMR contamination.

## Data Availability

All data produced in the present study are available upon reasonable request to the authors

## Acknowledgements

We thank Attila Fráter for assistance with the calibration and all participants for their time and commitment to this study. This work was supported by the Fonds Wetenschappelijk Onderzoek (FWO), grant G068621N (AuDiMod).

## Author Contributions

Conceptualization: SV, HK, PD; Methodology: PD, FD, SV, HK; Software: FD, PD; Investigation: PD; Formal analysis: PD, FD, MT; Visualization: PD; Writing – Original Draft: PD; Writing – Review & Editing: HK, SV, FD, MT; Supervision: SV, HK; Funding Acquisition: SV

## Statements and declarations

### Ethical considerations

This study was approved by the Ethics Committee of Ghent University (Ethics Code: ONZ-2022-0352 E01) on September 08, 2022. All participants provided written informed consent prior to enrolment in the study. This research was conducted ethically in accordance with the World Medical Association Declaration of Helsinki.

## Supplemental material

**Supplementary 1:**
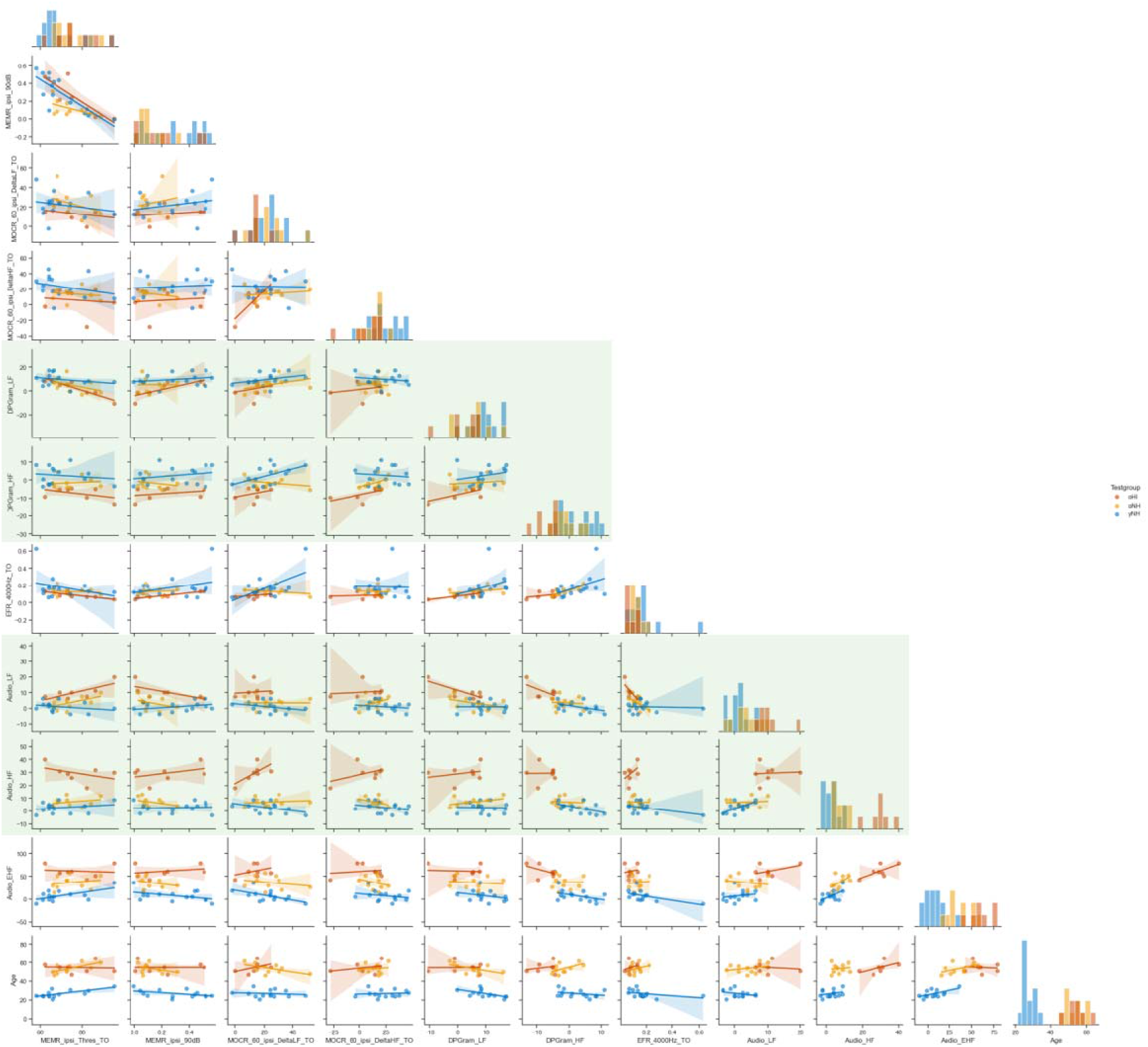
correlation matrices between the middle ear muscle reflex (MEMR) and the medial olivocochlear reflex (MOCR), and potential predictors for the hierarchical covariate adjustment analysis: distortion product otoacoustic emissions (DPOAE), the envelope-following response (EFR) and audiometry. The green boxes indicate the merged parameters, leaving four predictors: mean DPOAE, EFR at 4000 Hz, high-frequency audiometric thresholds, and extended high-frequency audiometric thresholds. LF = low-frequency; HF = high-frequency.

